# A Scoping Review of Artificial Intelligence Applications in Clinical Trial Risk Assessment

**DOI:** 10.1101/2025.01.21.25320310

**Authors:** Douglas Teodoro, Nona Naderi, Anthony Yazdani, Boya Zhang, Alban Bornet

## Abstract

Artificial intelligence (AI) is increasingly applied to clinical trial risk assessment, aiming to improve safety and efficiency. This scoping review analyzes 142 studies published between 2013 and 2024, focusing on safety (n=55), efficacy (n=46), and operational (n=45) risk prediction. AI techniques, including traditional machine learning, deep learning (e.g., graph neural networks, transformers), and causal machine learning, are used for tasks like adverse drug event prediction, treatment effect estimation, and phase transition prediction. These methods utilize diverse data sources, from molecular structures and clinical trial protocols to patient data and scientific publications. Recently, large language models (LLMs) have seen a surge in applications, representing over 20% of studies in 2023. While some models achieve high performance (AUROC up to 96%), challenges remain, including selection bias, limited prospective studies, and data quality issues. Despite these limitations, AI-based risk assessment holds substantial promise for transforming clinical trials, particularly through improved risk-based monitoring frameworks.

## Introduction

Clinical trials are key for evaluating the safety and efficacy of novel drugs and therapies^1,2^. Recent estimates of clinical trial failure show an increase for various phases and the average cost of a successful molecular entity has trended significantly upward for decades^3–5^. However, navigating the intricate landscape of potential risks associated with the execution of clinical trials can be a daunting^6^, nevertheless required task. Clinical trial risks encompass not only the well-being of participants (safety) and the ability of the intervention to deliver intended benefits (efficacy) but also the smooth and efficient execution of the trial itself up to drug approval (operational effectiveness)^4,7,8^. Identifying and mitigating these diverse risks throughout the trial process is crucial for improving patient safety as well as ensuring the integrity and ultimate success of the research enterprise.

Assessing risks in clinical trials presents several challenges both before and during the research process^9,10^. Firstly, ensuring adequate participant safety and informed consent remains paramount, requiring thorough evaluation of potential adverse effects and mitigation strategies^11^. Secondly, maintaining trial integrity amidst evolving scientific knowledge and external factors demands continuous monitoring and adjustment of risk assessments to uphold data validity and ethical standards^11,12^. Lastly, meeting regulatory and compliance obligations adds a layer of intricacy, necessitating careful adherence to protocols and guidelines to minimize legal and reputational risks while facilitating meaningful research outcomes^5^. Balancing these concerns is essential for effective risk management in clinical trials, safeguarding both participant well-being and scientific integrity throughout the research journey.

Research shows that artificial intelligence (AI) holds significant potential to enhance risk assessment in clinical trials^13^ by leveraging data-driven insights to improve safety, efficacy^14^, and operational effectiveness^15^ throughout the research lifecycle. AI algorithms can analyze vast amounts of data from various sources, including electronic health records, genomic data, and real-world evidence, to identify potential safety concerns, such as adverse events or drug interactions, more efficiently than traditional methods^16^. Additionally, AI-driven predictive models can assess the efficacy of interventions by analyzing complex patterns in patient data, aiding in the identification of promising treatments and patient subpopulations for targeted therapies^17^. Furthermore, AI-powered tools hold significant potential to streamline operational processes by optimizing trial design, patient recruitment, and data collection. These advancements increase the efficiency and cost-effectiveness of clinical research^18^.

Several reviews address the emerging potential and challenges of using AI in clinical trials from various perspectives. The works of Askin *et al.*^19^ and Harrer *et al.*^20^ offer comprehensive overviews of opportunities of AI for improved efficiency, recruitment, and faster trial design, while also acknowledging ethical concerns, data limitations, and regulatory hurdles. Weisller *et al.*^21^ take a multi-stakeholder approach, exploring machine learning applications across various trial phases. They acknowledge the need for more evidence and address ethical and philosophical barriers. Zame *et al.*^22^ focus on the specific challenges of health emergencies caused by pandemics. They demonstrate how machine learning can aid in predicting outcomes, repurposing drugs, and optimizing trial design in these urgent contexts. Other reviews, such as the works of Paul *et al.*^23^ and Patel and Shah^24^, either broadly discuss AI and machine learning in drug discovery, development, repurposing, clinical trials, and more, or focus on intervention safety^14^, such as the work of Basile *et al.*^25^. Finally, the review of Feijoo *et al.*^15^ analyzed specific operational risks within trials. Still, the literature lacks a scoping review highlighting the specific role of AI in the assessment of risks in clinical trials at large, particularly in light of recent advances in the field of machine learning. To bridge this gap, this work reviews the scope of the current literature on the application of AI methods in clinical trial risk assessment. More specifically, we aim to answer the following research questions:

1. What types of risks in clinical trials can AI methods be used to predict?
2. During which clinical trial phases and for which conditions are AI methods used to predict risks?
3. What are the dominant AI algorithms and data sources employed in clinical trial risk assessment?
4. How effective are AI methods in evaluating risks in clinical trials?
5. What are the key limitations of AI methods in predicting clinical trial risks?

The scoping review includes peer-reviewed articles published in English and was conducted in electronic databases, including PubMed, Web of Science, and Google Scholar. The review focuses on studies that apply AI algorithms to predict risks in clinical trials, including safety-, efficacy-, and operational-related risks, as in the framework proposed by Badwan *et al.*^14^. Overall, it provides a comprehensive overview of the current state of AI in clinical trial risk prediction and our findings could contribute to the development of more effective and efficient risk prediction strategies for clinical trials.

## Results

The initial search yielded 4’328 records from PubMed (n=1’605), Web of Science (n=1’086), and Google Scholar (n=1’628). After removing 1’026 duplicates, 3’302 records were evaluated for eligibility, with 3’108 being excluded and 2 not retrieved for full-text analyses. Of the remaining 192 records, 50 were excluded after full-text assessment, resulting in a final selection of 142 studies. The study selection flowchart is shown in Figure 1.

**Figure 1.**
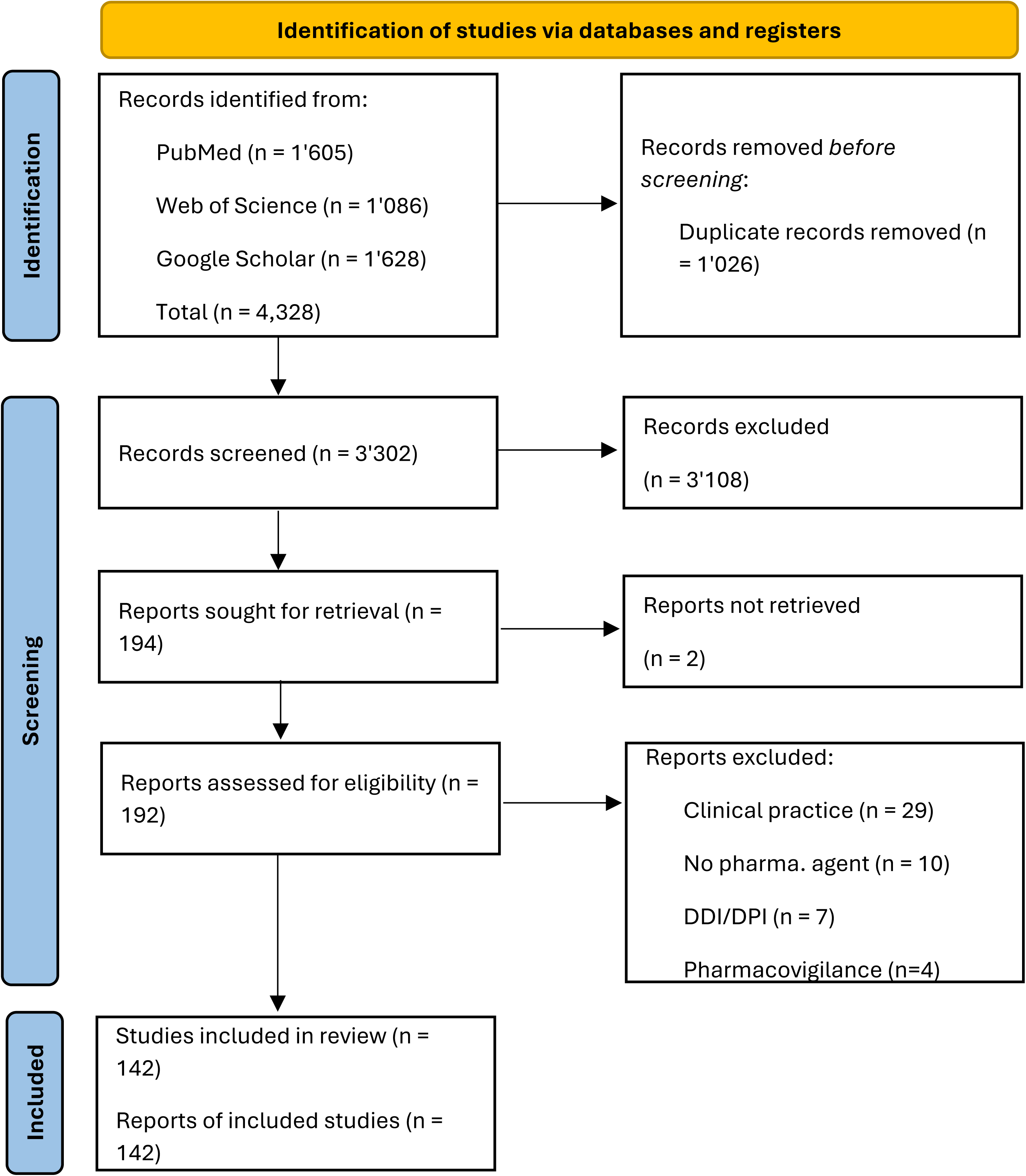
PRISMA flowchart describing the source of evidence retrieval and selection process. From the 4,328 manuscripts identified during the search phase, 3’302 titles and abstracts were screened after de-duplication, and 194 full texts. A total of 142 studies were included for full text analysis.

## What types of risks in clinical trials can AI methods be used to predict?

After the full-text analysis of the included articles, we categorized AI applications based on the type of clinical trial risk they address (Figure 2). To do so, we followed the framework proposed by Badwan *et al.*^14^, which describes three main phases in which AI methods can be used for risk prediction in clinical trials: toxicity, efficacy, and approval. In our analysis, we generalize the *toxicity* application to overall *safety* risk and consider *operational* risk instead of *approval* so that more fine-grained risk analysis can be taken into account, such as phase transition. An overview of the studies included in the review categorized by risk type is presented in Table 1.

**Figure 2.**
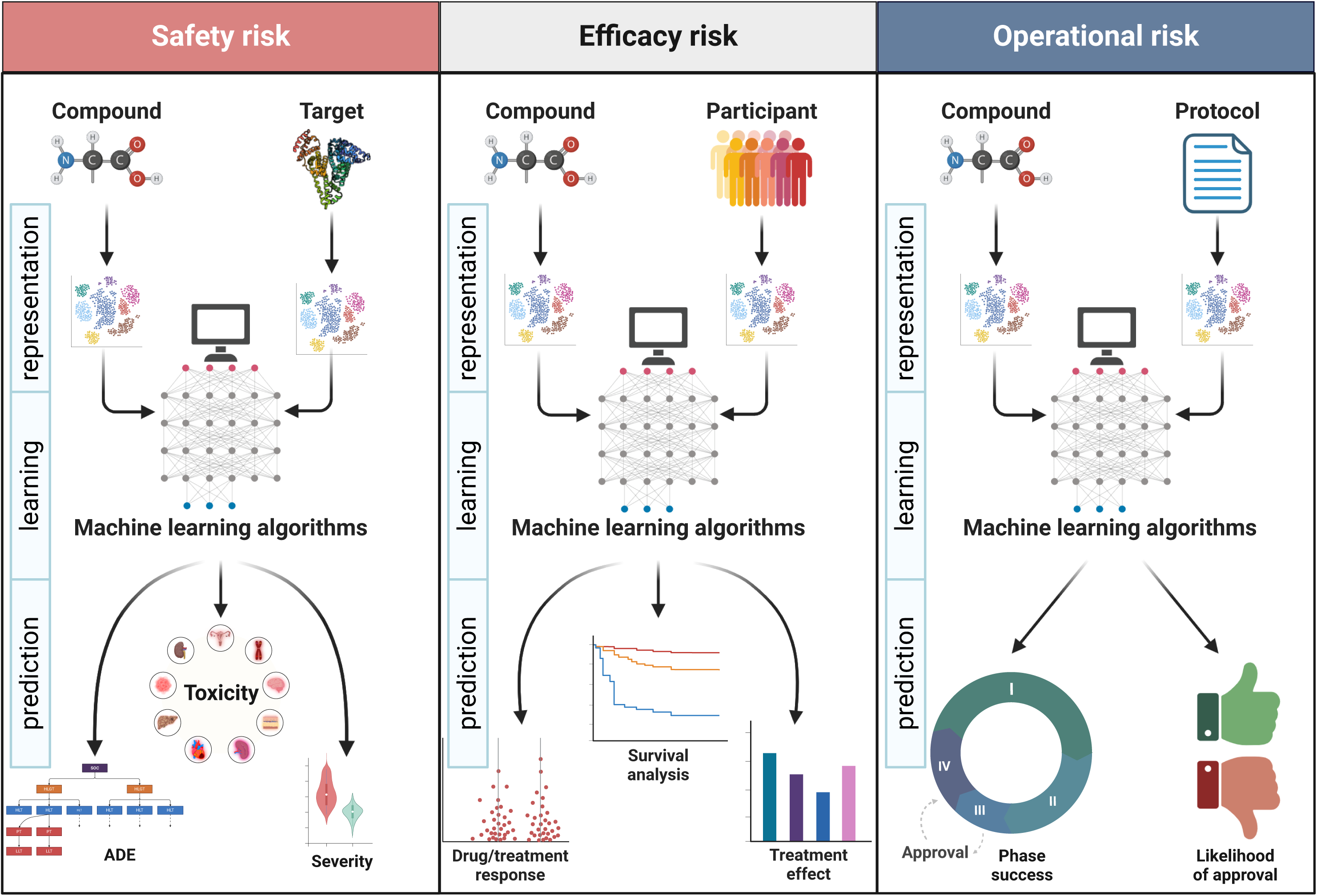
AI applications for risk assessment in clinical trials. AI applications can be categorized into safety, efficacy, and operational risk assessment. They follow a typical three-step analysis approach: representation, learning, and prediction (or inference). In the first step, clinical trial-related information, such as compound, participants, and protocol, are encoded as vectors. In the second step, models are learned to infer risks. In the last step, different risk types are predicted and performance metrics are obtained.

**Table 1.** Overview of the studies identified, classified by risk and application.

| Risk | Application | References | n |
| --- | --- | --- | --- |
| Safety | ADE | 26–43 | 18 |
| Safety | severity | 43–49 | 7 |
| Safety | toxicity | 50–80 | 32 |
| Efficacy | drug response | 83–93 | 11 |
| Efficacy | outcome | 94–106 | 13 |
| Efficacy | survival | 47,86,90,92,107–118 | 16 |
| Efficacy | treatment effect | 47,85,107–109,114,116,119–127 | 16 |
| Operational | likelihood of approval | 15,38,51,129–141 | 16 |
| Operational | phase success | 15,43,142–162 | 23 |
| Operational | other | 148,150,163–169 | 9 |

### Safety

As experimental treatments are being tested for the first time in humans, there may be unknown safety risks that are not fully understood until the trial is underway. The safety risk assessment category (Figure 2 - left) encompasses AI applications that, given a drug or compound, predict safety risks that can cause unexpected or severe adverse events to study participants, ranging from mild discomfort to serious health complications before the execution of a clinical trial^26–80^. Safety risk studies can be further subdivided into three predictive application use cases: *adverse drug event (ADE)*, *severity,* and *toxicity*. In *ADE* prediction^26–43^, AI models are designed to predict the occurrence of ADEs, that is, injuries resulting from the use of a drug^81^. These methods are usually multi-class, multi-label classifiers that infer the occurrence of adverse event categories, such as those proposed by MedDRA terminology^82^. Differently, methods for ADE *severity* prediction^43–49^ are usually binary classifiers that aim to infer the severity of ADEs, such as serious *vs.* non-serious or death *vs.* non-death events. Similarly, *toxicity* prediction methods^50–80^ are often binary classifiers that predict whether a drug or compound will be toxic for an organ, such as methods for predicting drug-induced liver injury (DILI)^51,56,59,61,63–68,70,73,79^.

### Efficacy

Efficacy risk assessment in clinical trials is the process of evaluating potential risks that could hinder the successful demonstration of a drug or treatment’s effectiveness (Figure 2 - center). AI applications within this category focus on predicting *drug response*, *outcome*, *survival*, and *treatment effect*^47,83–127^. In *drug response* applications, methods predict the potential for a drug to exhibit varying levels of efficacy across different patient populations or under specific conditions, often evaluated using in vitro models like cell lines to predict drug concentration and response^83–93^. In *outcome* prediction, the likelihood of a patient achieving a desired clinical outcome (e.g., disease remission, improved quality of life) following treatment is estimated^94–106^. These methods are typically assessed using binary classification tasks that predict the probability of response or non-response. In *survival* (or time-to-event) prediction, methods are specifically designed to analyze time-to-event data, such as time to death, disease progression, or recurrence^47,86,90,92,107–118^. They are particularly suited for risk prediction in the presence of censored data, where the event of interest might not have occurred for some individuals within the study period. Lastly, methods for *treatment effect* estimation^128^ quantify the differential impact of treatment on patient outcomes compared to a control group^47,85,107–109,114,116,119–127^. Unlike *survival* analysis, which explicitly models time-to-event data and competing risks, treatment effect estimation can be applied to a variety of outcomes, such as binary outcomes (e.g., disease remission *vs.* no remission) or continuous outcomes (e.g., blood pressure).

### Operational

Operational risk in clinical trials refers to the potential for disruptions, delays, or failures that can impact the successful execution and completion of a study, and ultimately the approval by regulatory agencies (Figure 2 - right). This category includes AI applications that assess the risk of the phase’s success, the likelihood of regulatory approval, or other operational factors, such as enrollment, duration, and site selection risk as well as informativeness of the protocol^15,38,43,51,129–169^. We categorized papers in the first subcategories following the approach of Feijoo *et al.*^15^. In the *likelihood of approval* risk, AI applications aim to estimate the overall probability of a drug receiving regulatory approval, often based on the study protocol^15,38,51,129–141^. In *phase success* prediction, AI applications are designed to estimate the probability of advancing a specific phase (e.g., from phase I to II)^15,43,142–162^. In practice, methods predict whether a clinical trial will complete the study graciously or terminate before completion. Lastly, we include in the *other* subcategory, the remaining operation risks, such as enrollment, duration, informativeness, etc., as their number of references was limited ^148,150,163–169^.

In Figure 3, we provide a high-level overview of the studies included in the analyses according to their publication date (Figure 3a), scientific subject area (Figure 3b), country of publication (Figure 3c), and type of publication venue (Figure 3d). We notice a growing interest in risk assessment of clinical trials based on AI, following an exponential growth trend (Figure 3a). The research in the field has an increasing trend since the past decade, with an important jump between 2020 and 2021. While between 2013 and 2020, the majority of studies focused on safety risk assessment (safety: n=18, efficacy+operational: n=18), from 2021 onwards there is a more even distribution in the three high-level risk categories (safety: n=37; efficacy: n=36; operational: n=37). Studies are published in multidisciplinary journals, in the fields of *Medicine*, *Computer Science* as well as *Biochemistry, Genetics and Molecular Biology*, which ranked in the top 3 subject areas of journals and conferences (Figure 3b). When considering the affiliation of the first author as the origin of the study, we notice a clear dominance of US institutions (n=53), followed by China (n=16) and the UK (n=10); South Korea (n=9) and Switzerland (n=7) complete the top 5 (Figure 3c). Lastly, 90% of the studies are published in peer-reviewed scientific journals, and the remaining 10% in peer-reviewed conferences (Figure 3d). The conference manuscripts were identified through Google Scholar searches. Out of the 14 manuscripts, three were presented at ICML workshops and three at EMNLP (n=2) and ACL (n=1) conferences. All the conference papers were classified within the "Computer Science" subject area but also included multidisciplinary conferences like the MLHC, which spans both "Computer Science" and "Medicine." Consequently, about one-third of the publications in the "Computer Science" category were presented at conferences.

**Figure 3.**
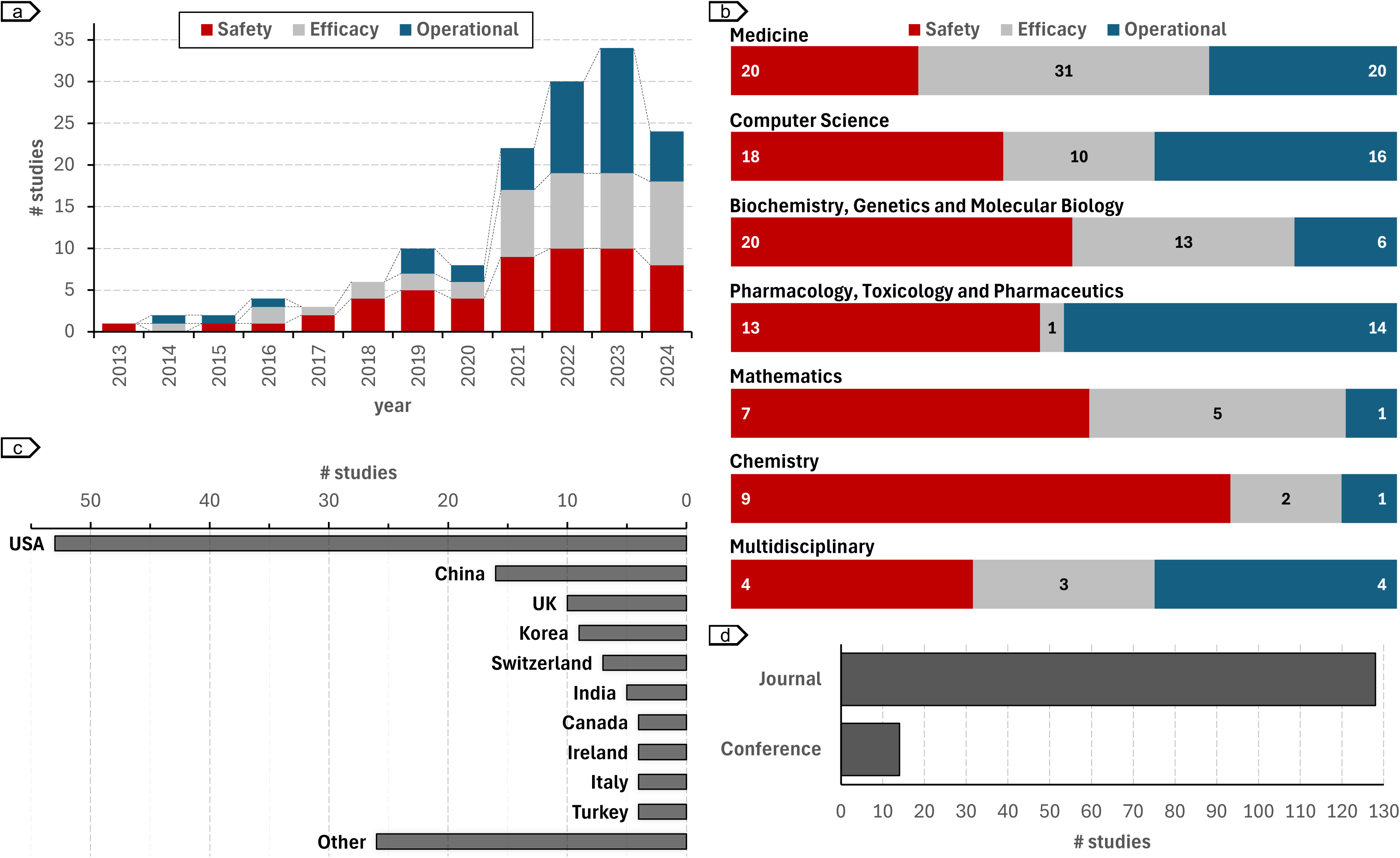
Trend of AI risk prediction for clinical trials. Distribution of manuscripts a) over time, and categorized by study b) subject area, c) country, and d) venue type.

## During which clinical trial phases and for which conditions are AI methods used to predict risks?

Since clinical trials face unique challenges depending on the phase and condition being investigated, we analyzed the distribution of studies based on these factors (Figure 4). Operational risk assessment methods are distributed across phases I-IV (Figure 4a) and tend to be phase-specific (n=37) (Figure 4b). In contrast, only a small fraction of safety risk assessment studies are phase-specific (n=10 *vs.* n=45 non-phase-specific), while efficacy risk studies are concentrated in phase III (n=21 *vs.* n=13 for phases I, II, and IV altogether). Regarding the condition studied in the clinical trial, most condition-specific risk assessment methods focused on *neoplasms* (n=29) followed by *mental disorders* (n=6) and *infections* (n=5) (Figure 4c). The majority of disease-specific studies focused on efficacy risk assessment (n=41 out 64). Figure 4d shows the distribution of adverse event-specific studies. All adverse event-specific studies are related to safety risk assessment (n=55). Studies predicting the risk of hepatic disorders (i.e., DILI) (n=16) together with those focused on multiple adverse events (n=16) represent 52% of the safety works.

**Figure 4.**
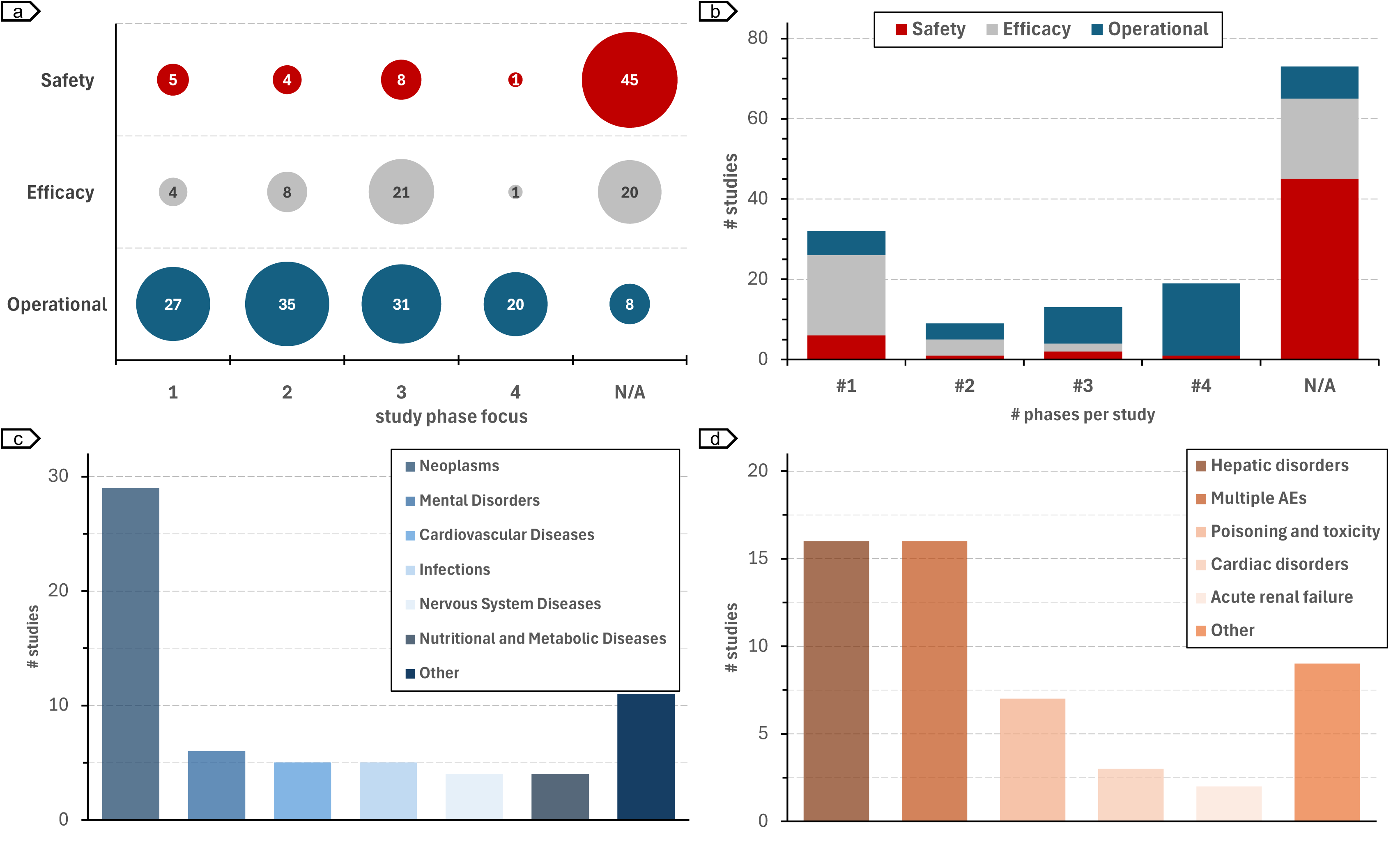
Distribution of studies according to phase, condition, and safety concerns. a) Number of studies focused on phases I-IV. b) Number of phases per study. c) Conditions in condition-specific studies. d) Safety categories in safety-specific studies.

## What specific AI methods and datasets are currently used to assess risks in clinical trials?

In Figure 5, we show the AI methods used for risk assessment in clinical trials. Based on the algorithm and prediction task, they can be categorized into six classes of machine learning analysis paradigms (a):

**Figure 5.**
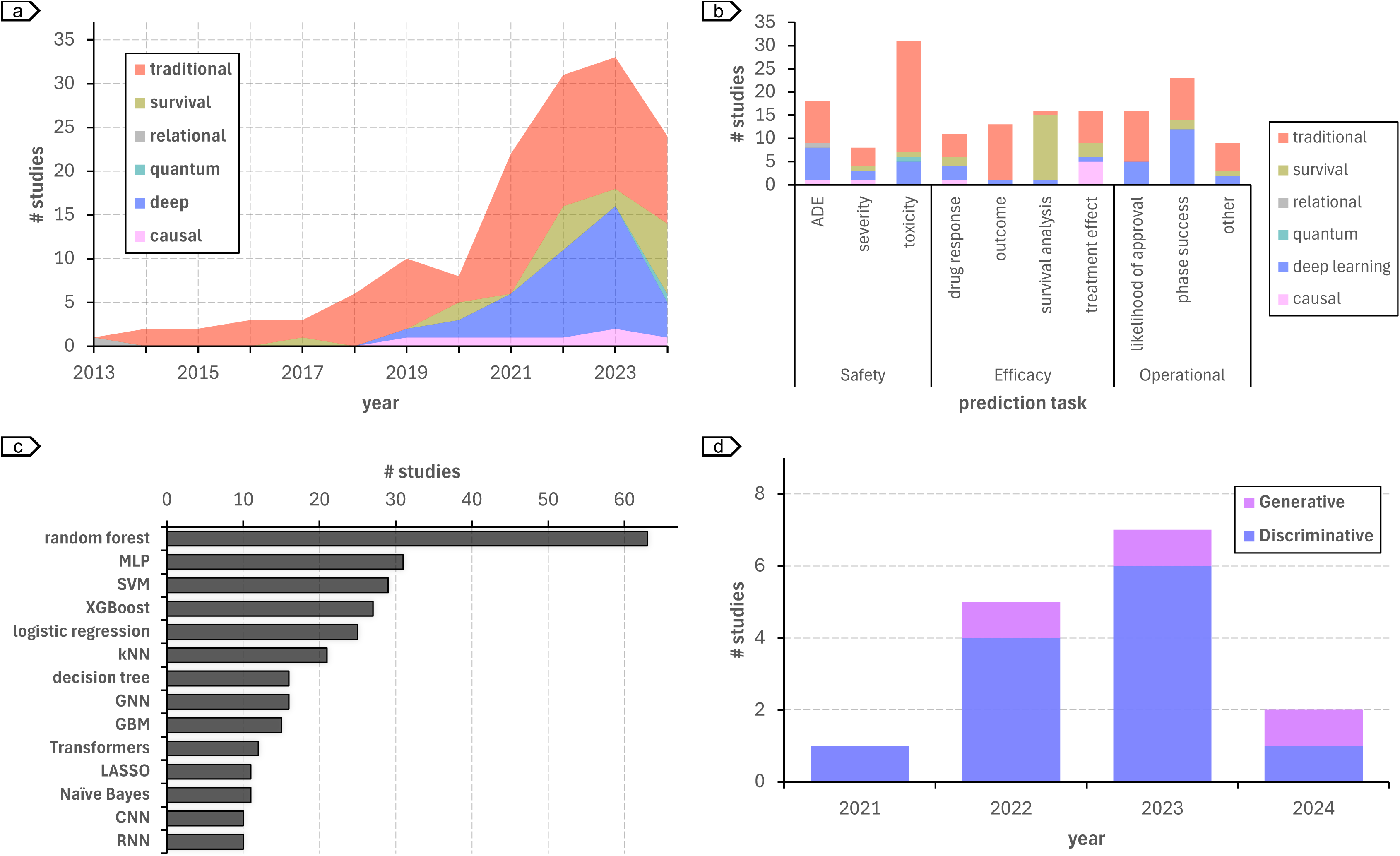
Machine learning models used in risk assessment of clinical trials. a) Trends of the different types of machine learning approaches for risk prediction of clinical trials. b) Approaches used for the different risk assessment tasks. c) Algorithms with results published in at least 10 risk assessment studies. d) Trends in utilizing large language models for clinical trial risk assessment.

**Traditional machine** learning includes algorithms that learn patterns from data to make predictions or decisions. These methods are often used with structured data extracted using feature engineering and are available from out-of-the-box toolkits, such as *scikit-learn* and *caret* in Python and R programming languages, respectively. As shown in a, traditional machine learning has been the dominant approach used in clinical trial risk assessment (safety: n=39; efficacy: n=28; operational: n=22). They are used across all the clinical trial risk prediction tasks (b). Examples of traditional machine learning algorithms used in clinical trial risk assessment include random forest, which is the most used algorithm (n=63), followed by support vector machines (SVM) (n=32), and extreme gradient boosting (XGBoost) (n=27) (c).

**Deep learning** is a subset of machine learning that uses artificial neural networks with multiple layers to learn complex patterns. Importantly, they can handle complex data types, such as free-text, images, graphs, etc., and extract features from the data automatically within a unified learning pipeline^170^. Together with traditional machine learning, deep learning is the main approach used in operation risk assessment (n=22) (b). Examples of deep learning algorithms used in clinical trial risk assessment include graph neural networks (GNNs) (n=16), transformers (n=12) convolutional neural networks (CNNs) (n=10), and recurrent neural networks (RNNs) (n=10) (c).

**Survival analysis** is a statistical learning method used to analyze time-to-event data, such as death or clinical trial termination, and includes methods such as the Cox proportional hazards model^171^. While survival analysis is often not considered a machine learning category in itself, we created a specific category for manuscripts including survival analyses as it accounts for time-to-event and for censoring data, which is an important feature in clinical trial risk analyses. Traditional and deep learning approaches can be combined with survival loss functions to provide survival curve predictions. Recent advances in deep representation learning have been expanded to include survival estimation, such as the DeepSurv model, which replaces the log-linear parameterization in classic models with a multi-layer perceptron^172^. Thus, approaches categorized into this category for clinical trial risk assessment^47,86,92,107–111,113–118,137,151,161,169^ can be based originally on traditional statistical learning as well as on deep learning. Most survival analysis works use traditional methods (n=12; the remaining n=6 are based on deep learning) and are applied to efficacy risk assessment (n=12) (b).

**Causal machine learning** is a specialized subset of AI that aims to identify cause-and-effect relationships between variables. This is distinct from traditional machine learning, which often focuses on identifying correlations. Causal models are used in clinical trial risk assessment to identify factors that directly influence a patient’s response to a treatment^47,87,108,116,120,125^. For instance, a causal model based on a multi-headed multi-layer perceptron (MLP) architecture was used for modeling the potential outcome of treatment and placebo on disability progression in multiple sclerosis^108^. This approach allowed researchers to estimate individual treatment effects and reduce the sample size required to detect an effect of the intervention. Together with survival analysis, causal inference is gaining traction in clinical trial risk assessment, particularly for treatment effect risk assessment (n=6) (b).

Two other AI approaches are used to predict risks in clinical trials: **relational learning** (n=1) and **quantum machine learning** (n=1). Relational learning is a classic AI technique that can handle data with complex relationships between entities. It includes methods such as inductive logic programming, which allows learning a concept definition from observations, i.e., sets of positive and negative examples, and background knowledge^173^. On the other hand, quantum machine learning is a nascent field that leverages quantum theory to model machine learning tasks. Both methodologies have been used to assess safety risks^26,78^. While relational learning did not increase in popularity, quantum machine learning has only recently been applied to assess clinical trial risks.

## The rise of LLMs

More recently, LLMs based on the transformer architecture have been successfully applied in clinical trial risk assessment^43,75,77,137,144,147,149,152,154,155,158,159,162^. In 2023, LLM-based studies represented already more than 20% (n=7) of the studies (d). On the other hand, the use of generative LLMs is still restricted, with only three studies between 2022 and July 2024. Ferdowsi *et al.*^144^ were the first to propose an AI method using LLM (encoder-based) to predict clinical trial risks. Their approach encoded sections of the clinical trial protocol using BERT, which were then fed into a GNN model to predict *phase success* risks. For safety risk assessment, the work of Morozov *et al.*^75^ used ProteinBERT to represent protein and predict toxicity. Katsimpras and Paliouras^137^ were one of the first to propose generative LLMs to predict clinical risk. Their approach was based on BART, an encoder-decoder model based on the transformer architecture. More recent examples of approaches using generative LLMs include inClinico^158^ and TWIN-GPT^43^. While the former used GPT-3.5 to extract clinical trial results from free-form text, such as publications and press releases, the latter explored the full power of LLM to create virtual trials and assess multiple risk types. To date, no approach based on LLM or transformers in general has been proposed in efficacy risk assessment.

## Data sources and performance metrics

Most studies assessing safety and operational risks used publicly available benchmarks to train and evaluate their models (safety: n=50; operational: n=37) (Figure 6a) while efficacy studies used mostly private datasets (n=32). These studies used datasets containing a median of 803 compounds, 1087 participants, and 17’538 protocols (Figure 6b), which are the main information types used for risk inference in safety, efficacy, and operational studies, respectively. Models were mostly evaluated using the area under the receiver operating characteristic (AUROC) curve (n=94), with accuracy (n=56) and recall (n=51) completing the top 3 metrics (Figure 6c). Only one metric in the top 10 reported, root mean square error (RMSE), refers to regression tasks. The remaining 9 metrics are used in classification tasks. Safety and operational studies reported a median of 3 metrics per study while efficacy studies reported 2 (Figure 6d).

**Figure 6.**
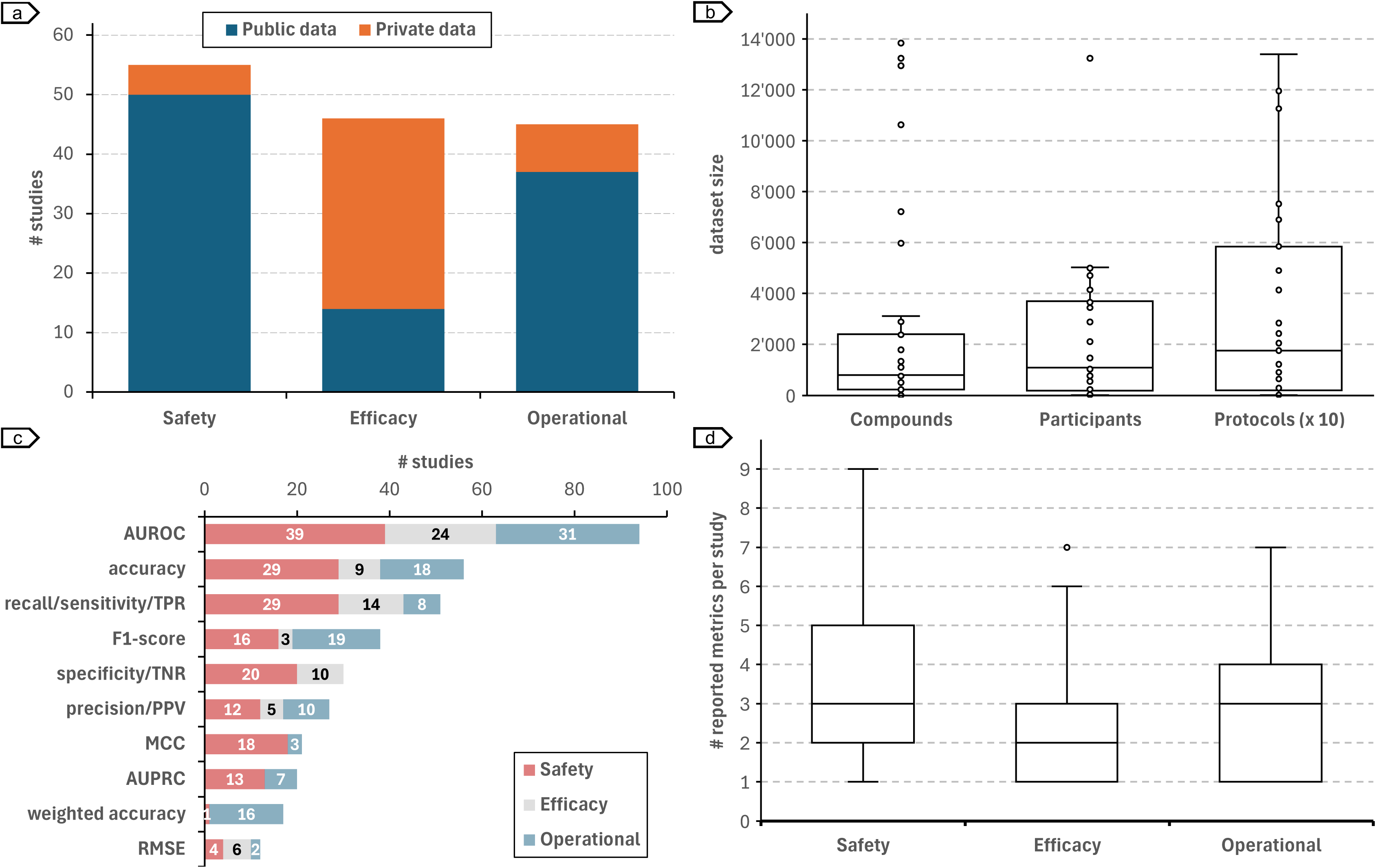
Datasets and metrics used to train and evaluate risk prediction models. a) Distribution of studies using (only) public and (at least one) private dataset. b) Number of compounds, participants, and protocols used to train and evaluate models. c) Top-10 metrics used in clinical trial risk assessment. d) Number of reported metrics per study.

Table 2 lists the most used datasets in safety, efficacy, and operational risk assessment studies. For safety risk, the main data source used was SIDER (Side Effect Resource)^174^, a comprehensive database of adverse drug reactions (ADRs) extracted from the FDA Adverse Event Reporting System (FAERS). Studies that used SIDER for safety risk assessment had a median number of 1’341 compounds in the benchmark dataset and the majority used only publicly available datasets (n=18). The main dataset used in efficacy studies was individual trials, with a medium number of only one trial and 1’250 participants per study. Most efficacy studies (n=29) used at least one private evaluation dataset. As for safety risk studies, operational risk used mainly public data sources (n=33), with most studies using data from clinicaltrials.gov.

**Table 2.** Main datasets used for safety, efficacy, and operational risk assessment studies.

| Risk | Data source | References | Instances (median) | Accessibility |  |
| --- | --- | --- | --- | --- | --- |
|  |  |  |  | Public | Private |
| Safety | SIDER | 26,28,30–36,38,39,42,55,57,60,70,71,77 | 1'341 compounds | 18 | 0 |
| Efficacy | individual trials | 47,85,87,89,92,94–96,98–107,107–110,112,114,115,117,119,121–127 | 1 clinical trial<br>1'250 participants | 5 | 29 |
| Operational | clinicaltrials.gov | 15,43,51,130,131,133,136,137,140–147,149–162,165–167 | 17'538 clinical trials | 33 | 3 |

## How effective are AI methods in evaluating risks in clinical trials?

Due to the different prediction tasks (Table 1), metrics (Figure 6c), and benchmark datasets (Table 2) used in clinical trial risk assessment, it is hard to compare the effectiveness of AI methods quantitatively. In an attempt to have a minimally comparative view, we analyzed performance across the largest subset of experiments using the same prediction tasks, reporting metrics, and dataset type. Figure 7a shows the performance across ADE (safety), outcome (efficacy), and phase success (operational) prediction tasks using SIDER, individual trial, and clinical trial protocol datasets, respectively, for studies reporting performance using the AUROC metric. In this subset, the top 3 *ADE* prediction studies all reported AUROC above 90%, with Masumshah *et al.*^30^ achieving the highest performance (96.6% AUROC), followed by Zhao *et al.*^31^ (93.1% AUROC), and Galeano *et al.*^28^ and Zhong *et al.*^42^ (92.0% AUROC). The top 3 *outcome* prediction models^100,103,104^ achieved AUROC between 84.0% and 87.4%, with Lei *et al.*^103^ achieving the best performance. For *phase success* prediction, Ferdowsi *et al.*^144,149,155^ rank in the top 3, with AUROC performance varying between 92.3% and 92.7%. For this subset of studies, the median number of instances in the datasets used in phase success prediction (n=75’174 protocols) is two orders of magnitude higher as compared to ADE (n=766 compounds) and outcome (n=235 participants) prediction experiments.

**Figure 7.**
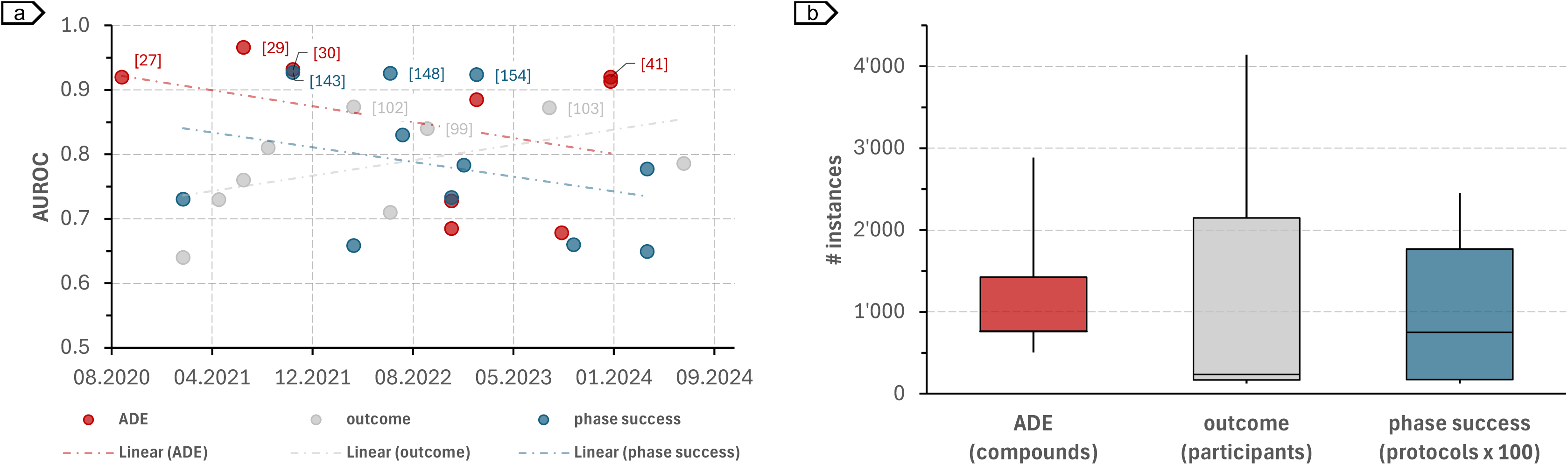
Model performance and dataset size. a) Model performance across ADE (safety), outcome (efficacy), and phase success (operational) risk assessment using SIDER, individual trial data, and clinical trial protocols, respectively. b) Number of instances used in the ADE, outcome, and phase success prediction tasks.

## What are the key limitations of AI methods in predicting clinical trial risks?

### Selection bias

Data used in clinical trial risk assessment might not be representative of the entire population of interest, leading to biased results. In safety risk studies, the number of compounds and drugs used in the experiment is significantly limited compared to the estimated drug-like chemical space (*O*(10^3^) vs. *O*(10^60^))^175^ or, more concretely, to the number of compounds in PubChem (*O*(10^3^) vs. *O*(10^8^)). Similarly, efficacy approaches are evaluated using individual trials, with a median number of trials per study equal to one (Table 2), that is, it is unclear if these methods will generalize to different interventions or conditions.

### Evaluation strategy

Datasets used in clinical trial risk assessment are often imbalanced. For example, for a given drug the number of negative ADE cases is far greater than the number of positive cases. Thus, results should be reported using metrics robust to class imbalance, such as F1-score, Matthew’s correlation coefficient (MCC), and weighted accuracy. However, as shown in Figure 6c, the top 3 most reported metrics are not robust to highly imbalanced datasets and could present high scores just by predicting the majority class.

### Data quality and availability

While operational risk studies tend to use large and representative sets of clinical trials, they often lack access to real-world clinical trial data, being mostly based on the design protocol (Table 2). Similarly, most safety risk studies ignore key factors, such as dosage and route of administration, when predicting toxicity and ADE risks.

### Retrospective studies

Despite some notable examples^103,158^, most clinical trial risk assessment studies use only retrospective data. While retrospective studies provide valuable insights into AI applications in clinical trial risk assessment, they may not be generalizable to different compounds, populations, or clinical trial contexts. This limits the applicability of the findings to broader clinical trial risk assessment.

### Siloed risk models

Clinical trial risks are assessed separately even though they are interconnected. A minimal dose (e.g., homeopathic) is unlikely to cause safety risks but also to statistically demonstrate efficacy against the condition under study. Similarly, interventions with severe safety risks might lead to operational trial termination. Despite these interdependences, only a few studies^38,43,47,51^, including TWIN-GPT^43^, consider a more holistic approach to clinical trial risk assessment. Nevertheless, they only investigate safety and a second category combined (efficacy^47^; operational^38,43,51^) and no study to date integrates the three risk assessment categories.

## Discussion

In this scoping review, we analyzed the existing literature on the applications of AI methods for predicting risks in clinical trials. Our review identified 142 studies describing safety (n=55), efficacy (n=46), and operational (n=45) risk assessment applications published in peer-reviewed journals and conferences between January 2013 and July 2024. We notice a growing interest in the field, with an exponential growth trend between 2013 and 2024. On the modeling methodology, various AI approaches have been used, including traditional machine learning, deep learning, and causal machine learning, in many risk prediction tasks, such as *ADE*, *treatment effect,* and *phase transition*. More recently, there was a surge in the applications of LLMs, reaching around 20% of the studies in 2023. However, the number of studies using generative approaches remains minimal. Models were trained and evaluated using a variety of benchmark datasets, both public, such as SIDER and clinicaltrials.gov, and private, such as individual trial data. The risk prediction models achieved high performance on some specific tasks, with AUROC as high as 96%. However, issues such as selection bias, poor evaluation strategies, and lack of prospective studies hinder the applications of the proposed methodologies in real-world scenarios. Nevertheless, AI-based risk assessment for clinical trials seems a promising research avenue and, based on the identified trends, is expected to grow in the coming years. In particular, these models could be used to improve risk-based monitoring frameworks in clinical trials and extend their adoption, which is not widespread^6^.

Four potential reasons contribute to drug development failure: unmanageable toxicity, poor drug-like properties, lack of clinical efficacy, and lack of commercial needs with poor strategic planning^176^. These failure reasons can be categorized into the three high-level risk types outlined in this review: safety, efficacy, and operational. AI-based safety methods aim to predict safety risks associated with the intervention, including potential adverse events, toxicity (e.g., DILI and drug-induced kidney injury), and severity (e.g., serious ADE and mortality). Efficacy methods predict whether an intervention is effective in treating a condition. Methods for efficacy risk assessment were further subdivided into approaches for predicting drug response and treatment outcome, estimating the treatment effect of an intervention, and progression-free survival. Only one study considered combined safety and efficacy risks^47^. Lastly, in operational risk assessment, AI-based methods were used to predict whether a drug will be approved, which depends on demonstrating its safety and efficacy, as well as whether a trial will complete a phase. It is important to note that completing a trial phase does not necessarily demonstrate the intervention’s safety (e.g., serious ADE events might have been identified) or efficacy (e.g., outcome measurements do not differ statistically between different interventions). Additionally, AI-based methods were used to predict issues related to the quality of the protocol and the trial process, such as recruiting. A few studies combined safety and operation risk prediction^38,43,51^. Despite their interconnection, risk prediction studies are rather siloed. A possible research avenue would be using multi-task learning for estimating multiple risk types at once, given that relevant data is available. For example, Yazdani *et al.*^177^ use a multi-task (or joint) learning approach to identify and normalize ADE-related entities in clinical notes. Similarly Tan^84^ uses multi-task learning to predict anti-cancer drug response. Multi-task learning methods typically combine the loss functions of different tasks to enhance data efficiency, reduce overfitting through shared representations, and accelerate learning by utilizing auxiliary information^84,84,178^. Thus, multi-task learning approaches could be employed to simultaneously and more effectively infer safety, efficacy, and operational risks.

The clinical trial process varies significantly according to the study phase. Thus, taking into account the phase but also experimenting across different phases is essential to capturing specific risks and generalizing to different scenarios. Interestingly, the majority of operational risk studies considered at least three phases (n=27 vs. n=18 for phases I, II, and non-phase-specific altogether), which could lead to better generalisability of such approaches as compared to the safety and efficacy AI-based risk assessment^42,105^, which are either non-phase-specific or focus on a single phase (Figure 4b). As the potential efficacy of compounds is already assessed in phase II, efficacy models focusing only on phase III, i.e., the majority, are likely to be biased. A notable example of efficacy risk study across phases I-III is described by Lu *et al.*^87^, proposing an application of deep learning to predict drug concentration and response time course. All the identified safety risk assessment studies are non-phase-specific. While ADEs are heavily dependent on the drug and dosage, the study population, which is phase-specific, has a non-negligible impact^82^. Additionally, an interesting application of in-silico risk assessment is to avoid providing potentially toxic interventions to participants. Thus, applications of AI risk could focus on phase I studies. In this line, the study of Bedon *et al.*^66^ proposes a machine learning model to predict the maximum tolerated dosage.

AI methods are applied in a variety of prediction tasks for clinical trial risk assessment, including i) *binary classification* as in toxicity (safety), treatment outcome (efficacy), and phase transition (operational) predictions, ii) *multi-class, multi-label classification* as in ADE prediction (safety), and iii) *regression* as in drug response and survival (efficacy) predictions. They use different types of input data, such as molecular structure, clinical information as well as semi-structure free-text clinical trial protocols. As a result, many types of AI algorithms based on machine learning techniques have been investigated. Safety prediction methods require methods for representing molecular structure but also relationships between drugs, genes, proteins, and ADEs. Thus, several studies leveraged the power of GNNs to capture these complex relationships and different modalities in an integrated machine learning pipeline^31,33,35,38,77^. Efficacy methods are often based on structured data derived from clinical trial results. As such, they tend to apply classic methods (including survival and causal learning) based on ensemble learning, such as random forest, gradient boosting machine (GBM), and XGBoost, for learning and inference^85,92,94–96,98–101,103,105–107,109,110,113,114,116,117,119,122–127^. Differently, in operational risk studies, the clinical trial protocol is often the focus of the analyses. Given the complex, semi-structured, free-text nature of the protocol, these methods tend to use deep learning, in particular, LLMs, to encode protocol information^43,137,144,147,149,152,154,155,158,159,162^. Indeed, apart from a few exceptions^43,137,158^, LLMs have been mostly used for textual representation (protocol, protein sequence, etc.) as dense vectors^77,144,155^. Natural language inference capabilities of LLMs have been little explored in this field.

We have identified some key limitations of existing AI-based clinical trial risk assessment studies. In particular, they include some selection biases and can have poor evaluation strategies, which can hinder their generalizability to different interventions or conditions and thus their real-world applications. Indeed, a recent study by Chekroud *et al.*^105^ found that machine learning accuracy for predicting patient outcomes is similar to chance when applied to out-of-sample trials. Despite the limitation of the study itself, which evaluates only one machine learning method (elastic net), being thus hardly generalizable to the ensemble of AI and machine learning methods, it gives some evidence of the limited generalizability of current AI studies for predicting efficacy risks. Moreover, for safety prediction, a recent study^42^ showed that advanced *ADE* prediction models do not differ significantly from a naïve classifier according to the AUROC metric, which is the main evaluation strategy used in this task. By only using the mean values of ADEs of known drugs to predict the ADEs for all the new drugs, the naïve model achieved 91% AUROC on the SIDER dataset, which is only 2 percentage points below the state-of-the-art model^31^.

Based on the identified limitations, several key recommendations can be made for future research in AI-based clinical trial risk assessment. First, data limitations for training and evaluating the models should be addressed. Researchers should increase the diversity and representativeness of datasets used in AI-based risk assessment, including data from real-world clinical trials and a wider range of compounds and outcomes, while ensuring high-quality data collection and annotation. For example, dosage and route of administration should be incorporated into safety risk assessment models to improve faithfulness to application scenarios, and large intervention-outcome scenarios should be considered. Moreover, evaluation strategies should be improved. Researchers should consistently employ evaluation metrics that are more robust to imbalanced datasets, such as F1-score, MCC, and weighted accuracy. Additionally, they should assess the generalizability of AI models to different interventions, conditions, and populations and on unseen data to demonstrate their real-world applicability. In this line, studies should move beyond retrospective analysis. Researchers should conduct prospective studies to establish causality and improve the generalizability of findings, as well as explore the use of real-time data, i.e., safety, efficacy and operational data collected during the study, to enable continuous monitoring and risk assessment during clinical trials. Finally, a promising research avenue would be to integrate risk assessment categories. In this direction, AI models could be developed to simultaneously predict multiple risk types, within the safety, efficacy, and operational categories. Thus, they would account for the interdependencies between different risk categories to provide a more holistic assessment.

This review has a few limitations. First, the field of AI is broad, which makes it challenging to identify the correct set of keywords for querying the databases. Some studies might focus on specific methodologies, e.g., SVM, XGBoost, and description logics, and do not mention in the abstract one of the high-level methodology-related keywords used in the search, i.e., AI, machine learning, etc. Thus, we might have missed some relevant studies. Despite that, we identified a large variety of modeling approaches, including even more classic AI methodologies, such as relational learning. Second, we took a strategic decision to guide our risk-related search keywords according to the application framework proposed by Badwan *et al.*^14^, which identified AI applications in clinical trials in three predictive areas: toxicity, efficacy, and approval. Instead, other viewpoints could be considered, for example, using components of risk-based quality management approaches^6^. This could be the subject of a specific and more targeted review. Finally, we were unable to fully answer one of the questions related to the effectiveness of the models for risk prediction due to the lack of common benchmarks and evaluation strategies used in the reviewed studies. We attempted to create a homogeneous set, focusing on specific tasks with common evaluation metrics and using the same benchmark datasets. Nevertheless, they are not comparable due to differences in the dataset used for training and evaluation, including size and content. Even so, these results provide a preliminary overview of the effectiveness of AI-based risk assessment in clinical trials.

In conclusion, this scoping review explored the potential of AI for risk assessment in clinical trials. We identified a rapidly growing field with diverse applications, focusing on safety, efficacy, and operational risks. AI models leverage various data sources, from molecular structures to clinical protocols, and employ techniques like traditional machine learning and, more recently, large language models. While some models achieve high performance, limitations exist. Selection bias and poor evaluation strategies hinder generalizability. Future research should address these issues by employing more diverse and representative datasets, incorporating real-world data, and focusing on generalizable evaluation metrics. Prospective studies and real-time (or continuous) data integration further hold promise. Additionally, exploring models that simultaneously predict multiple risk types could provide a more holistic assessment. Overall, AI holds significant promise for risk assessment in clinical trials, but further research is needed to ensure its real-world effectiveness.

## Methods

We conducted this systematic scoping review between October 2023 and July 2024. For processing and reporting the results of this review, we followed the guidelines of Preferred Reporting Items for Systematic Reviews and Meta-Analyses Extension for Scoping Reviews (PRISMA-ScR) guidelines^179^. The PRISMA-ScR checking list is provided in Supplementary Table 1.

## Search strategy and study selection

We collaborated with a health sciences librarian to develop a three-step search plan. Initially, we searched PubMed to locate key articles on risk assessment, clinical trials, and AI. These articles helped us create a comprehensive set of relevant keywords, organized into four categories: technology, application field, type of analyses, and task. We combined the keywords within each group using the OR operator and then linked all the groups using the AND operator. The search keywords used in the process are listed in Table 3. We then adjusted the syntax to ensure it could be used across the other databases. The systematic search was carried out in three databases – Medline, Web of Science, and Google Scholar – using their default search settings. Note that for the search in Google Scholar, we split Group 3 into three subgroups as the full query exceeded the character limit of the API. For each subquery, we considered for analysis the first 750 articles retrieved.

**Table 3.** Search keywords in different groups.

|  |  |
| --- | --- |
| Groupe 1 – methodology-related keywords | "artificial intelligence" OR "machine learning" OR "deep learning" OR "neural network" OR "text mining" OR "NLP" |
| Groupe 2 – study-related keywords | "clinical trial" OR "clinical study" OR "clinical trials" OR "clinical studies" |
| Groupe 3 – risk-related keywords | ("risk" OR "success" OR "termination" OR "completion" OR "outcome" OR "phase success" OR "likelihood of approval") OR ("adverse drug reaction" OR "adverse drug event" OR "adverse reaction" OR "adverse event" OR "adr" OR "ade" OR "toxicity" OR "safety") OR ("drug effectiveness" OR "efficacy") |
| Groupe 4 – task-related keywords | "predict" OR "prediction" OR "classify" OR "classification" OR "forecast" OR "forecasting" |
| Final result | (Groupe 1) AND (Group 2) AND (Group 3) AND (Group 4) |

## Eligibility criteria

In Table 4, we list the inclusion and exclusion criteria used in the screening and full-text analysis process:

- *Topic and methodology*: We considered studies on AI-based risk assessment in clinical trials. All forms of risk assessment, encompassing safety, efficacy, and operational considerations, were taken into account. Exclusions comprised healthcare applications, drug and protein interactions, and studies lacking pharmacological therapy. Studies encompassing predictive models for evaluating the risk of clinical trial interventions and/or the clinical study itself, based on AI, were incorporated. Information extraction studies, particularly those utilized in pharmacovigilance, along with standard risk factor analysis statistics, were excluded.
- *Context*: We have considered studies conducted across various geographic locations published in English. Following our initial screening, it became apparent that there were limited studies available on the AI-based risk assessment of clinical trials before 2013. Consequently, to concentrate on the most recent and pertinent AI-based risk assessment applications, we have restricted our focus to studies published from 2013 onwards.
- *Types of sources*: We included peer-reviewed studies from scientific journals and conferences. We included conference proceedings as important AI research is published in such venues. This also motivated the use of Google Scholar in our search, as it has a high literature coverage, including conferences. We excluded systematic and non-systematic reviews, dissertations, conference abstracts, observational studies, case reports, opinion pieces, commentaries, and protocols.

**Table 4.** Inclusion and exclusion criteria.

| Inclusion criteria | Exclusion criteria |
| --- | --- |
| Topic: Application of AI methods for clinical trial risk assessment | Topic: Healthcare application (as opposed to clinical research) |
| Language: English | Topic: Drug and protein interaction |
| Publication date: from 01.01.2013 to 15.07.2024 | Topic: Non pharmacological therapy (radiotherapy, gene therapy, food, etc.) |
| Review process: Peer reviewed articles | Methodology: Risk factor analyses (e.g., using logistic regression) and other non-predictive methods in general (e.g., information extraction). |
| Article type: Basic research |  |
| All articles retrieved in PubMed and WoS, and the first 750 articles in Google Scholar for the 3 Google Scholar queries |  |

## Dataset screening

We used Bibdesk to de-duplicate the results, sorting articles by key, title, and first author. Identified duplicated articles were manually checked. Then, we uploaded all the results to an online spreadsheet to make it easier for the team to review and extract information from titles, abstracts, and full texts. To ensure a convergent approach, two reviewers pilot-tested the screening of titles and abstracts on a random sample of 10 studies. After the pilot test, two independent reviewers examined each article. If they disagreed on whether to include a citation, a third reviewer would evaluate the citation, and we would then discuss it together to reach a consensus. We uploaded the full texts of potentially relevant articles to Zotero for further screening by two reviewers. Any differences in including or excluding full-text studies were resolved during a consensus meeting.

## Dataset annotation

We randomly divided the articles included for full-text review among four researchers. Each researcher read the full-texts separately and used a standard spreadsheet to extract item information based on the CHARMS (Critical Appraisal and Data Extraction for Systematic Reviews of Prediction Modelling Studies) checklist^180^. The spreadsheet included study characteristics (authors, publication date, country of first author, subject area, journal or conference), and information about the source of data (data source and public availability), outcome to be predicted (type of risk analysis, prediction task, and ADE), candidate predictors (study phase and condition, compound, clinical trial data, and clinical trial protocol), sample size (dataset size and dataset type), model development (algorithm, model type, and training strategy), model performance (evaluation metrics), and results (performance on the test set). We piloted the template, added a "not available" option for some items, and included examples to enhance consistency and ease of use. After extraction, an independent reviewer normalized and consolidated the information.

## Data analyses

We analyzed the data using Microsoft Excel for Mac (Microsoft Office 365, version Version 16.89.1). We used descriptive statistics like frequencies, median, and ranges, and presented the data graphically and in tabular format as needed. We summarized the study characteristics, including the frequency and distribution of publication year, country of the first author, subject area, and type of publication venue. Additionally, we examined dataset and evaluation characteristics, such as mean/median size, frequency/distribution of phase, condition, ADE, and metrics, as well as model considerations, including the frequency/distribution of algorithms and training strategies, and performance.

## Data Availability

The data underlying this article can be shared on reasonable request to the corresponding author.

## Data Availability

The data underlying this article can be shared on reasonable request to the corresponding author.

## Acknowledgements

This research received no specific grant from any funding agency in the public, commercial, or not-for-profit sectors.

## Author Contributions

D.T. conceptualized the study, defined the methodology, performed the database searches and coordinated the screening process. D.T. also performed analyzed the data and authored the original draft. N.N. and D.T. performed the screening process. N.N., A.Y., B.Z., and A.B. extracted item information from full-texts. All authors reviewed and approved the manuscript.

## Competing Interests

The authors declare no competing interests.

## Notes

### Competing Interest Statement

The authors have declared no competing interest.

### Funding Statement

This study did not receive any funding.

